# Exploring the Genetic Link Between Thyroid Dysfunction and Common Psychiatric Disorders: a Specific Hormonal, or a General Autoimmune Comorbidity

**DOI:** 10.1101/2022.05.17.22275202

**Authors:** Sourena Soheili-Nezhad, Emma Sprooten, Indira Tendolkar, Marco Medici

## Abstract

**Background:** The hypothalamus-pituitary-thyroid axis coordinates brain development and post-developmental function. Thyroid hormone variations, even within the normal range, have been associated with the risk of developing common psychiatric disorders, although the underlying mechanisms remain poorly understood.

**Materials and methods:** To get new insight into the potentially shared mechanisms underlying thyroid dysfunction and psychiatric disorders, we performed a comprehensive analysis of multiple phenotypic and genotypic databases. We investigated the relationship of thyroid disorders with depression, bipolar disorder, and anxiety disorders in 502,480 subjects from UK Biobank. We subsequently investigated genetic correlations between thyroid disorders, thyroid stimulating hormone (TSH) and free T4 (FT4) levels, with the genome-wide factors that predispose to psychiatric disorders. Finally, the observed global genetic correlations were furthermore pinpointed to specific local genomic regions.

**Results:** Hypothyroidism was positively associated with an increased risk of major depressive disorder (OR=1.51, p<10^−16^) and bipolar disorder (OR=1.99, p=2.1×10^−6^). Genetically, strong coheritability was observed between autoimmune hypothyroidism and both major depressive (r_g_=0.17, p=2.7×10^−4^) and anxiety disorders (r_g_=0.17, p=6.7×10^−6^). This genetic correlation was particularly strong at the Major Histocompatibility Complex (MHC) locus on chromosome six (p<10^−5^), but further analysis showed that other parts of the genome also contributed to this global effect. Importantly, neither TSH nor FT4 levels were genetically correlated with mood disorders.

**Conclusion:** Our findings highlight an underlying association between autoimmune hypothyroidism and mood disorders, which is not mediated via thyroid hormones, and in which autoimmunity plays a prominent role. While these findings could shed new light on the potential ineffectiveness of treating (minor) variations in thyroid function in psychiatric disorders, further research is needed to identify the exact underlying molecular mechanisms.

## Introduction

Thyroid hormone (TH) plays a key role in brain development and neurocognitive function [1, 2]. Both hypo-and hyperthyroidism have been associated with an increased risk of various psychiatric diseases [3], including depression, bipolar and anxiety disorders [4]. More recently, it has even been shown that minor subclinical variations in thyroid function, even within the normal range, are associated with these psychiatric diseases [5, 6], although some studies report contradicting findings [7, 8]. A better understanding of the biological processes underlying these associations could have highly valuable implications for both clinical psychiatry and endocrinology.

The most common forms of thyroid dysfunction have an autoimmune origin and there is increasing evidence that the immune system and inflammatory response are also relevant for the pathophysiology of psychiatric disorders [9, 10]. Next to the direct effects of TH on brain, the effects of thyroid dysfunction on psychiatric diseases could thus be mediated by more generally associated autoimmune processes [11]. Various cellular mechanisms have been proposed for this phenomenon, ranging from binding of thyroid peroxidase antibodies (TPOabs) to human astrocytes, coinciding anti-central nervous system auto-antibodies in thyroid diseases, to increased production of monocyte- and T-lymphocyte derived cytokines [11]. Vice versa, psychiatric disorders have also been shown to be associated with abnormalities in thyroid function tests [12, 13]. These include mild elevations as well as decreases in thyroid hormone levels, blunting of the TSH response to TRH stimulation, and absence of the nocturnal surge of TSH. In addition, psychiatric disorders have also been suggested to be associated to altered autoimmune processes [14].

Taken together, there is still a large knowledge gap whether the observed associations between (minor or overt) thyroid abnormalities and psychiatric diseases are directly mediated via TH or not. This is a clinically relevant gap, as understanding the complex interactions between thyroid status, autoimmunity, and psychiatric disorders is a prerequisite before making treatment recommendations to prevent or treat coinciding psychiatric disorders in a patient with mild thyroid dysfunction, or vice versa.

In the context of these complex interactions, it is noteworthy that an estimated 60-70% of the inter-individual variation in thyroid function is determined by genetic factors [15]. Next to a direct genetic relationship between thyroid function and psychiatric disorders, various studies have suggested that genetic factors play an important role in various other autoimmune diseases as well as psychiatric disorders [16–18]. In recent years, genome-wide association studies (GWAS) have been particularly successful in identifying the genetic determinants of thyroid function [19, 20], and more recently of psychiatric disorders [21]. With the availability of these data, this is the optimal moment to investigate the role of common genetic variants in the complex relations between thyroid function, autoimmunity and common psychiatric diseases.

Doing so could help understand whether the well-known clinical comorbidity between thyroid diseases and psychiatric disorders is specifically driven by thyroid hormones, or has a more general autoimmune-related basis. Therefore, in the current study, we tested phenotypic and genetic correlations in large datasets of thyroid diseases, thyroid hormone levels, and three common psychiatric disorders coinciding with thyroid dysfunction, i.e., major depressive disorder, bipolar disorder and anxiety disorder.

## Methods

### Phenotypic correlation

We studied the co-occurrence of thyroid dysfunction and psychiatric disorders in 502,480 subjects of UK biobank. The clinical diagnosis of hyperthyroidism or hypothyroidism was recorded as International Classification of Diseases (ICD-10) codes (table S1). Subjects with the history of both hypo- and hyperthyroidism were removed from the analysis (n=1,178). In the current work, we considered three common psychiatric disorders, including major depressive disorder (MDD), bipolar disorder (BIP) and anxiety disorder (ANX). These phenotypes were extracted from various data fields and questionnaires similar to previous studies (table S2). The statistical association of thyroid disease with each of the three psychiatric disorders was assessed using logistic regression while controlling for the confounding effects of sex, age, age^2^, and the interactions between sex×age and sex×age^2^.

### Global genetic correlation

The genetic correlations of TSH and FT4 and thyroid diseases with ANX, MDD and BIP were estimated using LD-score regression (https://github.com/bulik/ldsc [22, 23]) and the largest GWAS summary statistics available for each phenotype at the date of analysis: the GWAS of TSH and FT4 levels within the normal range [24], TSH in the entire range [25] (i.e., including unaffected subjects and subjects affected by thyroid disease), a recent GWAS of auto-immune thyroid disorders [26], bipolar disorder [27], anxiety disorder [28] and major depressive disorder [29] as available at the psychiatric genomics consortium portal (https://www.med.unc.edu/pgc [30]). GWAS data were filtered using minor allele frequency >0.01 and an imputation info score cut-off of 0.9. The GWASs of multiple phenotypes in UK biobank have been previously performed by Neale lab and generously released for third-party use. This dataset includes the GWAS of thousands of phenotypes related to lifestyle, general health and medical history conditions (https://github.com/Nealelab/UK_Biobank_GWAS). We retrieved the SNP summary statistics of self-reported Hypothyroidism and Hypothyroidism phenotypes from this repository (table 1).

**Table 1.**
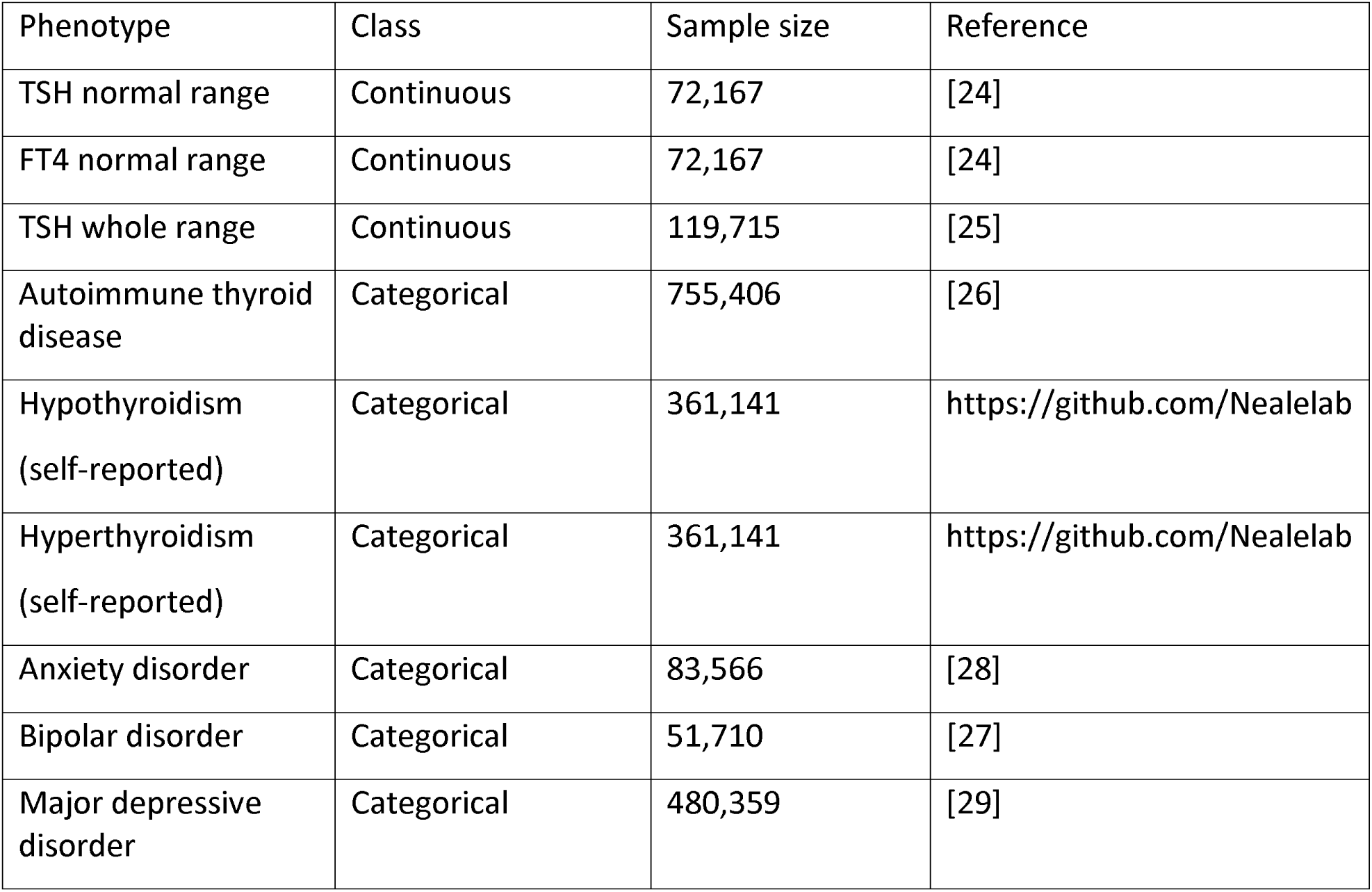
GWAS summary data used in the current work

### Local genetic correlation

For those phenotype pairs demonstrating significant global genetic correlations, we subsequently tested their local genetic correlation using Local Analysis of Variant Annotation (LAVA [31]). This procedure divides the genome into 2,495 local linkage disequilibrium (LD) blocks and then estimates genetic correlations within each LD block.

After identifying LD blocks of strong genetic correlation using LAVA, these “hotspots” were removed from the global genetic correlation analyses, to estimate the remaining genetic correlation excluding the local hotspots. This analysis was aimed at understanding whether the coheritability of thyroid disease and psychiatric disorders was driven by a small number of strong local genetic correlations or was rather distributed at many loci across the genome.

## Results

### Phenotypic correlations

First, we verified associations between thyroid dysfunction, mood and anxiety disorders in a large population-based cohort (UK Biobank), the results of which are shown in Table 2. The most common mood disorder was major depressive disorder (broad definition [29], with a prevalence of 35.1% in the cohort), followed by anxiety disorder (5.3%) and bipolar disorder (0.2%). The prevalences of hypothyroidism and hyperthyroidism were 3.8% and 0.3%, respectively. Hypothyroidism was positively associated with an increased risk of major depressive disorder and bipolar disorder, while hyperthyroidism was associated with an increased risk of major depressive disorder (table 2). The association of hypothyroidism with bipolar disorder was particularly strong (OR=1.99). No significant phenotypic correlations were observed between either thyroid disease and anxiety disorder.

**Table 2.** Phenotypic associations between thyroid dysfunction and three psychiatric disorders

|  | No anxiety disorder (%) | anxiety disorder (%) | Total (row-wise, %) | OR | z-score (p) |
| --- | --- | --- | --- | --- | --- |
| Normal thyroid function | 453,223 (94.7) | 25,129 (5.3) | 478,352 (96.0%) |  |  |
| Hypothyroidism | 17,653 (94.1) | 1,110 (5.9) | 18,763 (3.8%) | 1.05 | 1.50 (0.133) |
| Hyperthyroidism | 1,309 (93.3) | 94 (6.7) | 1,403 (0.3%) | 1.17 | 1.50 (0.134) |
| Total (column-wise) | 472,185 (94.7) | 26,333 (5.3) | 498,518 |  |  |
|  | No bipolar disorder (%) | Bipolar disorder (%) | Total (row-wise) | OR | z-score (p) |
| Normal thyroid function | 477,574 (99.8) | 778 (0.2) | 478,352 |  |  |
| Hypothyroidism | 18,711 (99.7) | 52 (0.3) | 18,763 | 1.99 | 4.74 (2.16×10<sup>-6</sup>) |
| Hyperthyroidism | 1,402 (99.93) | 1 (0.07) | 1,403 | 0.457 | -0.78 (0.434) |
| Total (column-wise) | 497,687 (99.8) | 831 (0.2) | 498,518 |  |  |
|  | No major depressive disorder (%) | Major depressive disorder (%) | Total (row-wise) | OR | z-score (p) |
| Normal thyroid function | 312,767 (65.4) | 165,585 (34.6) | 478,352 |  |  |
| Hypothyroidism | 9,817<br>(52.3) | 8,946<br>(47.7) | 18,763 | 1.51 | 27.09<br>($<10^{-16}$) |
| Hyperthyroidism | 805<br>(57.4) | 598<br>(42.6) | 1,403 | 1.24 | 3.86<br>($1.11 \times 10^{-4}$) |
| Total (column-wise) | 323,389<br>(64.9) | 175,129<br>(35.1) | 498,518 |  |  |

### Global genetic correlations

Next, we investigated global genetic correlations between thyroid traits and mood and anxiety disorders, the results of which are shown in Table 3, along with the observed SNP-heritability estimates (h2-SNP). Among the traits, hyperthyroidism had very low SNP heritability (0.006), and was therefore not included in subsequent analyses.

**Table 3.** Global genetic correlation between thyroid hormone levels and thyroid diseases with affective disorders

| | h<sup>2</sup> SNP | Anxiety disorder $r_g$ (p) | Major depressive disorder $r_g$ (p) | Bipolar disorder $r_g$ (p) |
| --- | --- | --- | --- | --- |
| h<sup>2</sup> SNP |  | 0.08 | 0.07 | 0.35 |
| Hypothyroidism | 0.05 | 0.17 ($2.2 \times 10^{-4}$) | 0.17 ($8.5 \times 10^{-5}$) | 0.02 (0.52) |
| TSH normal range | 0.11 | 0.05 (0.43) | -0.05 (0.37) | -0.04 (0.42) |
| TSH entire range | 0.10 | 0.02 (0.67) | -0.03 (0.55) | -0.06 (0.12) |
| FT4 normal range | 0.15 | -0.02 (0.71) | -0.04 (0.36) | -0.03 (0.55) |
| Autoimmune thyroid disease | 0.03 | 0.16 ($6.7 \times 10^{-6}$) | 0.13 ($2.7 \times 10^{-4}$) | 0.004 (0.89) |

We observed strong genetic correlations between autoimmune thyroid diseases and major depressive disorder (r_g_=0.13, p=2.7×10^−4^) and anxiety disorder (r_g_=0.16, p=6.7×10^−6^), but not with bipolar disorder. As autoimmune thyroid diseases include both autoimmune hypothyroidism as well as Graves’ disease, the results were validated using an independent GWAS on hypothyroidism in UK biobank (table 3), concluding that the observed associations were predominantly driven by autoimmune hypothyroidism.

Next, to explore whether the observed genetic correlations with hypothyroidism are driven by thyroid hormone levels or autoimmunity (i.e., independently of thyroid hormone levels), genetic correlations between thyroid hormone levels, mood and anxiety disorders were tested. Despite higher SNP-heritability estimates for TSH and FT4, there were no significant genetic correlations between TSH or FT4, either in the normal or the entire range, and any of the mood and anxiety disorders.

### Local genetic correlation

Finally, we carried out local genetic correlation analyses to refine the genomic regions underlying the observed global genetic correlations between hypothyroidism, mood and anxiety disorders. A strong local genetic correlation block was observed between autoimmune thyroid disease, anxiety and major depressive disorders at loci 6p22.1 and 6p22.2 (table 4 and 5), flanking the major histocompatibility complex (MHC) region. Another such “hotspot” of local genetic correlation was identified in chromosome 12, in a region containing multiple genes (RAP1B, NUP107, SLC35E3, MDM2, CPM, CPSF6, LYZ, YEATS4, FRS2, CCT2, LRRC10, BEST3; table 4 and 5).

**Table 4.** Top local genetic correlations between autoimmune thyroiditis and anxiety disorder

| Locus | Chr | start | stop | SNP count | PCs | Rho | p |
| --- | --- | --- | --- | --- | --- | --- | --- |
| 952 | 6 | 27261036 | 28666364 | 316 | 41 | 0,87 | $6.5 \times 10^{-6}$ |
| 1804 | 12 | 68839662 | 70097805 | 459 | 135 | 1 | 0,00011 |
| 951 | 6 | 26396201 | 27261035 | 202 | 32 | 0,72 | 0,00060 |
| 267 | 2 | 59251997 | 60775066 | 626 | 178 | -0,67 | 0,00071 |
| 932 | 6 | 6621938 | 7808323 | 519 | 179 | -0,81 | 0,00077 |
| 2072 | 15 | 69089816 | 70767983 | 486 | 166 | 0,80 | 0,0015 |
| 2453 | 21 | 39252060 | 40480831 | 511 | 175 | -0,69 | 0,0021 |
| 861 | 5 | 10290398<br>6 | 103788460 | 271 | 89 | -0,77 | 0,0032 |
| 1559 | 10 | 79952997 | 81190573 | 529 | 172 | -0,61 | 0,0035 |
| 1215 | 7 | 13875510<br>8 | 140217630 | 325 | 117 | -0,75 | 0,0057 |
| 2485 | 22 | 41722788 | 42867927 | 277 | 59 | 0,84 | 0,0065 |
| 1391 | 9 | 15839404 | 16658377 | 413 | 141 | -0,70 | 0,0097 |
| 1671 | 11 | 60515106 | 61717117 | 269 | 94 | 0,71 | 0,012 |
| 226 | 2 | 22429641 | 23538527 | 315 | 102 | -0,86 | 0,012 |
| 203 | 2 | 1376546 | 1875060 | 186 | 72 | -0,72 | 0,013 |
| 1576 | 10 | 98005995 | 99367800 | 466 | 125 | -0,75 | 0,014 |
| 2022 | 14 | 95946793 | 97174314 | 539 | 165 | -1 | 0,016 |
| 1059 | 6 | 13145716<br>5 | 132890694 | 404 | 123 | 0,61 | 0,016 |
| 35 | 1 | 36855140 | 38474036 | 514 | 137 | 0,73 | 0,018 |

**Table 5.**
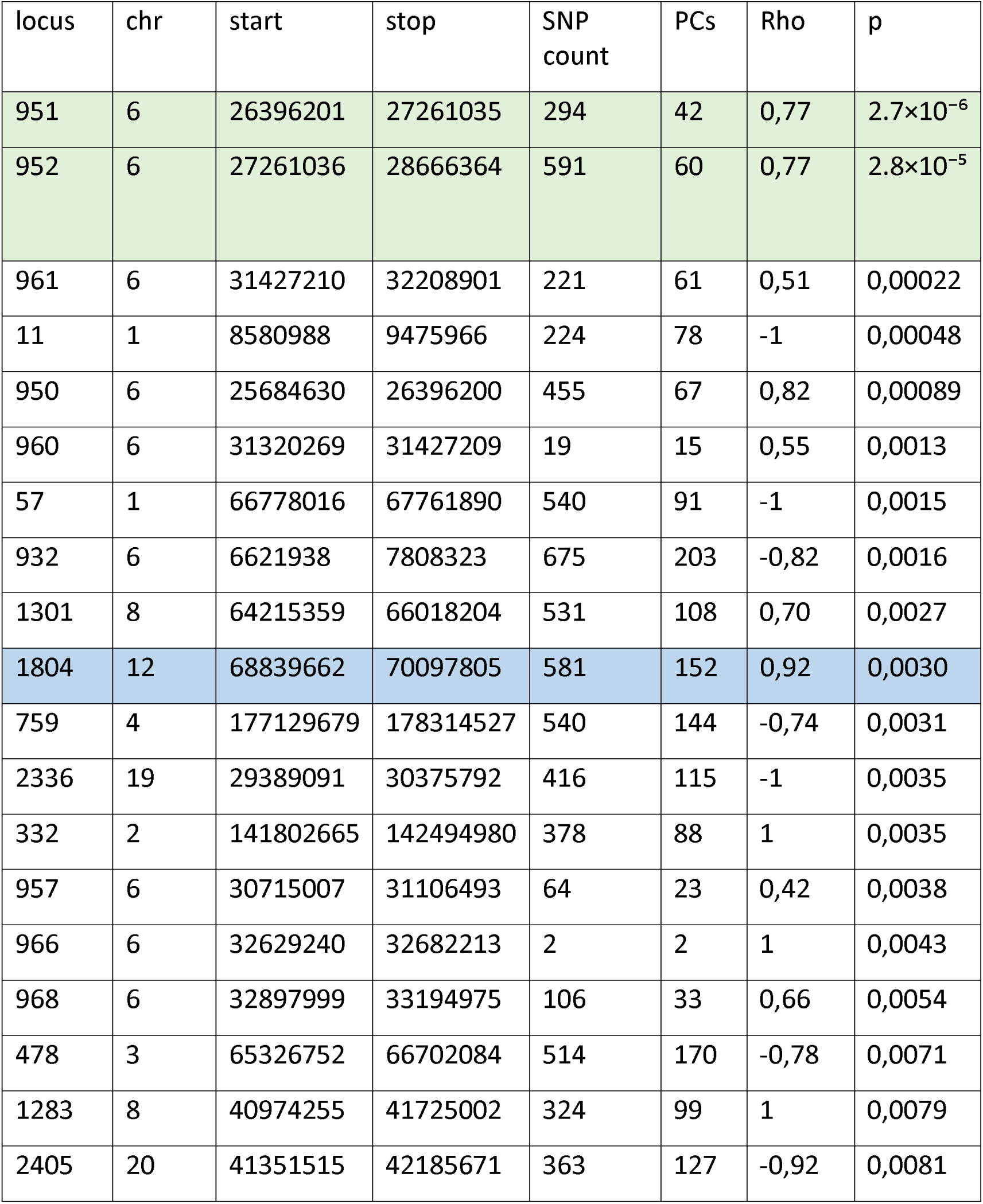
Top local genetic correlations between autoimmune thyroiditis and major depressive disorder

| locus | chr | start | stop | SNP count | PCs | Rho | p |
| --- | --- | --- | --- | --- | --- | --- | --- |
| 951 | 6 | 26396201 | 27261035 | 294 | 42 | 0,77 | $2.7 \times 10^{-6}$ |
| 952 | 6 | 27261036 | 28666364 | 591 | 60 | 0,77 | $2.8 \times 10^{-5}$ |
| 961 | 6 | 31427210 | 32208901 | 221 | 61 | 0,51 | 0,00022 |
| 11 | 1 | 8580988 | 9475966 | 224 | 78 | -1 | 0,00048 |
| 950 | 6 | 25684630 | 26396200 | 455 | 67 | 0,82 | 0,00089 |
| 960 | 6 | 31320269 | 31427209 | 19 | 15 | 0,55 | 0,0013 |
| 57 | 1 | 66778016 | 67761890 | 540 | 91 | -1 | 0,0015 |
| 932 | 6 | 6621938 | 7808323 | 675 | 203 | -0,82 | 0,0016 |
| 1301 | 8 | 64215359 | 66018204 | 531 | 108 | 0,70 | 0,0027 |
| 1804 | 12 | 68839662 | 70097805 | 581 | 152 | 0,92 | 0,0030 |
| 759 | 4 | 177129679 | 178314527 | 540 | 144 | -0,74 | 0,0031 |
| 2336 | 19 | 29389091 | 30375792 | 416 | 115 | -1 | 0,0035 |
| 332 | 2 | 141802665 | 142494980 | 378 | 88 | 1 | 0,0035 |
| 957 | 6 | 30715007 | 31106493 | 64 | 23 | 0,42 | 0,0038 |
| 966 | 6 | 32629240 | 32682213 | 2 | 2 | 1 | 0,0043 |
| 968 | 6 | 32897999 | 33194975 | 106 | 33 | 0,66 | 0,0054 |
| 478 | 3 | 65326752 | 66702084 | 514 | 170 | -0,78 | 0,0071 |
| 1283 | 8 | 40974255 | 41725002 | 324 | 99 | 1 | 0,0079 |
| 2405 | 20 | 41351515 | 42185671 | 363 | 127 | -0,92 | 0,0081 |

Contrary to our expectation, after excluding these “hotspots” of local genetic correlation, the calculated global genetic correlation remained high (anxiety disorder p=4.2×10^−6^ and major depressive disorder p=2.7×10^−4^), indicating that the observed genetic correlations are spread across the genome rather than localized to a few hotspots.

## Discussion

In the current study, we systematically analyzed the role of common genetic variants in the complex relations between thyroid function, autoimmunity and three common psychiatric disorders. We found strong genetic correlations between autoimmune hypothyroidism, major depressive disorder and anxiety disorders. Despite higher SNP-heritability estimates for thyroid function (TSH and FT4), there were no significant genetic correlations between TSH and FT4 and any of these psychiatric disorders. This may suggest that the genetically determined relation between hypothyroidism and mood/anxiety disorders is not mediated via thyroid hormones directly, but via pathways that underpin disease pathogenesis, such as (general) autoimmunity. This was further supported by our regional genetic correlation analyses, in which the top locus was the MHC region, which plays a central role in the pathogenesis of various autoimmune disorders, including autoimmune hypothyroidism [32]. It has been known for long that inflammatory pathways play a central role in the pathogenesis of autoimmune hypothyroidism. However, in the last decade various studies have shown that inflammatory pathways also play a key role in the pathogenesis of many psychiatric disorders [33–35]. Notably, studies on inflammatory biomarkers have been unable to detect markers that are specific for any particular psychiatric disorder, thereby suggesting genetic variation in autoimmunity to be a more general underlying phenomenon. Various biological treatment regimes, including pharmacotherapy and non-invasive brain stimulation with electroconvulsive therapy (ECT) indeed lead to a reduction of pro-inflammatory markers, highlighting the role of inflammation in psychiatric vulnerability [36, 37]. Next to inflammatory pathways leading to both autoimmune hypothyroidism and increased psychiatric vulnerability, coinciding autoimmune disorders might also for other reasons than shared inflammatory pathways be responsible for the observed relations between thyroid and psychiatric disorders. Indeed, it has been shown that autoimmune hypothyroidism coincides with several other autoimmune conditions, such as rheumatoid arthritis, celiac disease, type 1 diabetes, premature ovarian failure and adrenal insufficiency [38]. These comorbidities have also been associated with various psychiatric diseases, including MDD and anxiety disorders. Therefore, future studies in well-phenotyped cohorts should assess which part of the observed associations between hypothyroidism and psychiatric disorders is actually driven by these other autoimmune comorbidities. Furthermore, one of the several mechanistic hypotheses underlying the coincidence of multiple autoimmune conditions is that various tissues share the same epitopes, to which autoantibodies can cross-react [39]. In this context, Benvenga et al. [40] recently showed an interesting amino acid sequence homology between thyroid autoantigens and central nervous system proteins, including thyroid peroxidase, autoantibodies to which are a characteristic biomarker for Hashimoto’s hypothyroidism.

Irrespective of the exact underlying mechanisms, our findings have also clinical relevance. Based on the well-known clinical correlations, thyroid dysfunction is among the first somatic disorders to be ruled out in a patient with a newly diagnosed psychiatric disorder. Various studies have investigated whether correction of milder forms of thyroid dysfunction (including subclinical hypothyroidism) improves depressive and anxiety-related symptoms, with disappointing results [41, 42]. Our finding that the genetically determined relation between hypothyroidism and mood/anxiety disorders is not mediated via thyroid hormones, but more likely by (general) autoimmunity, is a plausible explanation for this lack of effects, and supports a restrictive attitude towards treating mild thyroid dysfunction in psychiatric patients.

Besides psychiatric disorders, milder affective and anxiety related complaints are frequently reported in hypothyroid patients. Unfortunately, these complaints persist in 10-15% of levothyroxine treated patients despite normalization of their thyroid hormone levels, which is among the largest knowledge gaps in endocrinology [43]. Our findings suggest that the psychiatric vulnerability in Hashimoto’s hypothyroidism patients is at least partly mediated via autoimmunity and not thyroid function, which would be a plausible explanation as to why these patients have persisting complaints despite the normalization of their thyroid hormone levels.

Our combined phenotypic and genetic analyses across multiple psychiatric and thyroid traits allowed us to better understand their shared biological underpinnings, more so than we would have in a purely phenotypic analysis. While our evidence for genetic overlap strongly encourages further research into shared molecular mechanisms, the approaches we used remain observational and do not directly exclude alternative causal explanations. For example, heritable lifestyle factors (such as diet and physical activity) are shared predisposing factors to both psychiatric disorders and thyroid diseases, so these could be contributing mediating factors in addition to any directly mediating, purely molecular inflammation pathways [44–48]. Future research using mendelian randomisation [49] and longitudinal designs are warranted to better inform on the different possible causal mechanisms.

Our local genetic correlation analysis identified two loci as potential mediators of the genetic overlap between thyroid and mood/anxiety disorders. Notably, one of these two regions maps to 6p22.1. Common variants in this region have been previously implicated in schizophrenia [50]. This locus flanks the major histocompatibility complex (MHC) genes and is a genetic correlation ‘hub’ across multiple, mostly (auto)immune-related clinical conditions [31].

Our genetic analysis was made possible by the recent surge in well powered GWAS data from very large sample sizes and consortia, both in psychiatry and endocrinology. The resultant power, and the fact that our genetic correlations are based on independent samples, strengthen the present study. However, a limitation of using existing GWAS statistics is that they differ in statistical power, and consequently the genetic correlations are not directly comparable across traits. Despite being based on larger sample sizes than the autoimmune hypothyroidism analyses, no genetic correlation was found between TSH/FT4 and psychiatric disorders. This strengthens the finding that the observed association between autoimmune hypothyroidism and psychiatric disorders is for an important part autoimmune related.

It is difficult to know whether the strong phenotypic correlation we found between bipolar disorder and hypothyroidism is not driven by genetic variation at all, or whether it is due to lower statistical power in the bipolar disorder GWAS data we used. Nevertheless, GWAS data for major depressive disorder and schizophrenia were very well powered, and provided robust results that were consistent across two independent GWASs of thyroid disease. Similarly, differences in prevalence across the different diseases is a limitation of the phenotypic correlation analysis, especially in a population-based sample like UK Biobank. Especially for hyperthyroidism, this limits the conclusions we can draw. In contrast, the higher prevalence for hypothyroidism and major depression led to well powered phenotypic correlation analyses and conclusive results.

In conclusion, our results indicate that autoimmunity rather than thyroid hormone itself plays a key role in the genetically determined relations between hypothyroidism and mood/anxiety disorders. Next to providing valuable clinical insights into the potential benefits of treating milder forms of thyroid dysfunction, these findings warrant further research into the autoimmune mechanisms underpinning these common coexisting diseases that may lead to a shift in personalizing treatment in the future.

## Data Availability

All data produced in the present work are contained in the manuscript.

## Supplementary Tables

**Table S1.** Thyroid disease definition based on ICD diagnosis codes

|  | ICD-10 codes | #Subjects |
| --- | --- | --- |
| Hypothyroidism | E039 | 20,904 |
|  | E063 | 301 |
| Hyperthyroidism | E050 | 681 |
|  | E052 | 140 |
|  | E059 | 2,105 |
| Exclusion codes | C73, D34, D093, E051, E053, E054, E058, E060, E061, E062, E064, E065, E069, E030, E031, E032, E033, E034, E035, E038, E890 | 3,028 |

**Table S2.** Psychiatric disorder definition based on UK biobank data fields

| Disorder | Field numbers | #Subjects |
| --- | --- | --- |
| Anxiety disorder (ANX) | Field 20544: mental health problems ever diagnosed by a professional (codes 1,5,6,15 and 17)<br>-Social anxiety or social phobia<br>-Any other phobia (e.g., disabling fear of heights or spiders)<br>-Panic attacks<br>-Anxiety, nerves or generalized anxiety disorder<br>-Agoraphobia<br>-no comorbidity: schizophrenia, ADHD, ASD, BIP, Bulimia nervosa, psychological over-eating or binge-eating | 26,333 |
| Major depressive disorder (MDD) | Fields 2090 and 2100<br>-Seen doctor (GP) for nerves, anxiety, tension or depression<br>-Seen a psychiatrist for nerves, anxiety, tension or depression<br>Field 41202: diagnoses - main ICD10<br>-F32: Depressive episode<br>-F33: Recurrent depressive disorder<br>-F34: Persistent mood [affective] disorders<br>-F38: Other mood [affective] disorders<br>-F39: Unspecified mood [affective] disorder | 175,129 |
| Bipolar disorder (BIP) | Field 20544: mental health problems ever diagnosed by a professional (code 10)<br>-Mania, hypomania, bipolar or manic-depression | 831 |

## Notes

This work was supported by funding from the Radboud University (Healthy Brain Study pre-seed grant).

### Competing Interest Statement

The authors have declared no competing interest.

### Author Declarations

The study used only openly available human genome-wide association study summary statistics data that were originally located at various online repositories, including the Thyroidomics consortium portal (https://transfer.sysepi.medizin.uni-greifswald.de/thyroidomics/datasets/), psychiatric genomics consortium (https://www.med.unc.edu/pgc), UK biobank GWAS portal by Neale lab (https://github.com/Nealelab/UK_Biobank_GWAS) and deCODE (https://www.decode.com/summarydata).

